# Change in profile of COVID-19 deaths in the Western Cape during the fourth wave

**DOI:** 10.1101/2022.01.12.22269138

**Authors:** Masudah Paleker, Mary-Ann Davies, Peter Raubenheimer, Jonathan Naude, Andrew Boulle, Hannah Hussey

## Abstract

Fewer COVID-19 deaths have been reported in this fourth wave, with clinicians reporting less admissions due to severe COVID-19 pneumonia when compared to previous waves. We therefore aimed to rapidly compare the profile of deaths in wave 4 with wave 3 using routinely collected data on admissions to public sector hospitals in the Western Cape province of South Africa. Findings show that there have been fewer COVID-19 pneumonia deaths in the Omicron-driven fourth wave compared to the third wave, which confirms anecdotal reports and lower bulk oxygen consumption by hospitals in the province.

## Background

Increasing evidence suggests that South Africa’s fourth wave of SARS-CoV-2 infections, caused by the Omicron variant, is associated with less severe disease [1–3]. A lower absolute number of COVID-19-related deaths have been reported in this fourth wave, and anecdotally clinicians have reported more “incidental” COVID-19 in hospitalized patients, and fewer patients requiring oxygen support for COVID-19 pneumonia. We aimed to rapidly compare COVID-19 deaths in the third (Delta driven) wave vs. the fourth wave in the Western Cape.

## Methods

We conducted a descriptive cross-sectional analysis using routine electronically available data collated by the Western Cape Provincial Health Data Centre. The first 50 deaths among patients admitted to public sector hospitals with SARS-CoV-2 infection from when there were 5/million admissions in wave 3 (15 June 2021) and wave 4 (6 December 2021) were assessed.

We included deaths during admission or within 14 days of discharge from a public sector hospital in the Western Cape. Each deceased case was independently investigated by two clinicians (HH and MP) using the Single Patient Viewer [4] by reviewing the available linked data, including laboratory tests, medication prescribed, ICD-10 coding, and electronic clinical records (emergency triage data and discharge summaries), to determine the likely cause of death. Deaths were categorised as follows:

- Severe COVID-19: evidence of COVID-19 pneumonia
- No COVID-19 pneumonia:
- COVID-associated: no evidence of COVID-19 pneumonia and presence of another medical condition as the primary cause of death e.g., diabetic keto-acidosis, stroke, active tuberculosis or malignancies and COVID-19
- Incidental: asymptomatic COVID-19 and other cause of death, where COVID-19 did not precipitate admission or death (e.g., trauma, surgery), or explicitly documented as incidental by attending clinician
- Indeterminate: insufficient information, or cannot exclude other pathology as being primary cause of death

Differences in categorization between the reviewers were resolved by consensus. Ethics approval was obtained from the Human Research Ethics Committee, University of Cape Town (460/2020).

## Results

50 deaths occurred within 5 days in wave 3 and 12 days in wave 4. In the fourth wave, 50% of COVID-19 deaths occurred in those with evidence of COVID-19 pneumonia, compared to 78% in the third wave (see Figure 1), while 24% and 2% of COVID-19 deaths were deemed COVID-associated in the fourth and third waves respectively, with the most common primary diagnoses in COVID-19 associated deaths in wave 4 being malignancy (10%) and active tuberculosis (8%).

**Figure 1:**
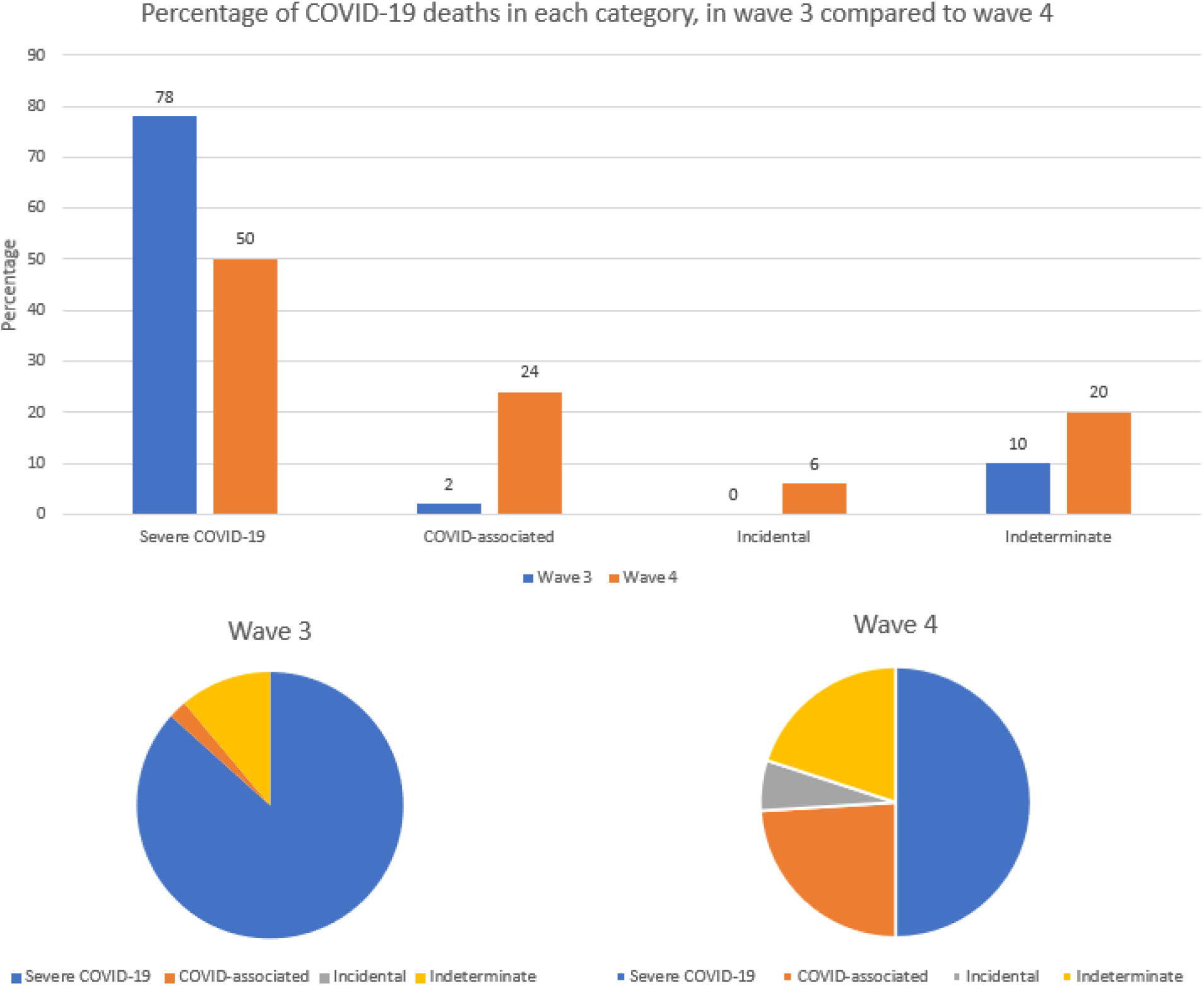
Bar graph showing percentage of COVID-19 deaths in each category in wave 3 compared to wave 4, with pie charts below showing the breakdown per wave.

## Conclusion

These findings concur with anecdotal reports from clinicians, and lower health facility bulk oxygen consumption in this fourth wave [5]. While there is a smaller proportion of COVID-19 pneumonia in wave 4, there are increasing numbers of COVID-associated deaths where COVID-19 is not presenting with pneumonia but may have precipitated acute admission in already sick patients or may be completely incidental.

While most health authorities will continue to report “COVID-19 deaths” as all deaths in patients with a positive COVID-19 test, these need to be interpreted appropriately in the context of increasing SARS-CoV-2 endemicity and immune-escape, widespread testing of admitted and deceased patients and high SARS-CoV-2 prevalence at wave peaks.

## Data Availability

All data produced in the present study are available upon reasonable request to the authors

